# Review of Teleradiology Applications in Some Selected African Countries

**DOI:** 10.1101/2023.10.07.23296691

**Authors:** Jacob Nii Noye Nortey, Linda Akorfa Agbotsigah, Eric Opoku Osei, Andrew Adabo, Miriam Gborgblah, Rashida Suleiman

**Affiliations:** Kwame Nkrumah University of Science and Technology, Department of Computer Science

**Keywords:** Telemedicine, Teleradiology, Digital Health, Ghana, South Africa, Developing Countries

## Abstract

The role of radiology is central to disease management however, its application is hindered by the low distribution of radiology worldwide with the situation direr in Africa. Information and communication technology (ICT) methods have been used to s healthcare systems and lessen the negative after-effects of widespread unavailability of high-quality radiological services. Telemedicine, specifically teleradiology was identified as a prime mitigator resulting in increased access to quality radiological services and effective diagnostic reporting thus contributing to achieving universal health coverage. Around the world, teleradiology adoption and implementation has seen significant boost and healthcare benefits but in Africa, the situation is reversed. Africa’s low adoption can be linked to policymakers not having adequate evidence-based data to inform decisions and policies on teleradiology implementation. In this study, we review the implementation of teleradiology in five (5) African countries: South Africa, Egypt, Morocco, Kenya, and Ghana. Articles were searched on Databases such as PubMed, Google Scholar, and ScienceDirect using defined search strings. Sixty papers were initially downloaded, but only those meeting the inclusion criteria were retained after the titles and abstracts were reviewed. Nineteen articles were found suitable for the study after careful assessment. All of these were summed up, and the study’s most important takeaways were plotted. Few studies met our inclusion criteria, indicating that teleradiology is not widely used in the nations we looked at. As a result, it is necessary to conduct extensive investigations to direct the widespread implementation of teleradiology in Africa. The findings show that teleradiology is been implemented in these countries however, there exists the challenge of lack of technology, policies, human sources, and training that are hindering teleradiology practices. Therefore, it recommended that various governments and scholars should address the challenges facing teleradiology and examine the limitations identified in this study.

## 1.1 Introduction

Telehealth is a broad concept that covers the technologies which provide administrative and clinical long-distance support for patients, physicians, and other health-related persons through the use of information and telecommunication technologies and services such as video conferencing, store-and-forward imaging, monitoring, and other non-clinical services, [1]. Telemedicine, on the other hand, is the provision of high-quality healthcare services through the use of communications technologies to facilitate remote exchanges of patient information, [2]. Non-clinical services like tele-education and tele-training, and clinical services like telenursing, teleradiology, tele-pharmacy, telecardiology, tele-laboratory, etc. are also provided over vast distances, [1]. This is a new phrase for describing our increasingly interconnected global society, which has profound implications for how we think about and approach health and medical care, [3].

Teleradiology is a branch of telemedicine whereby telecommunication systems are used to transmit, view and interpret diagnostic or radiological images from one location to another for diagnostic or consultative purposes by a radiologist or physician, [4]. Examples of such imaging techniques include X-ray imaging, Computed Tomography (CT), and Magnetic Resonance Imaging (MRI), among others. Teleradiology enables radiologists to deliver services without being present at the patient’s location(s), [5]. The reason for its increased implementation is that teleradiology addresses the lack of adequate professionals to provide radiological coverage as well as the general lack of expertise in this field of specialization, [6].

In Ghana, for instance where there are fifty-six (56) reported radiologists serving the 31 million population with almost all concentrated in the urban settlements, [7]. A patient in a rural settlement in Ghana will have to travel several kilometers to an urban health facility to access a radiologist for image interpretation which is impractical in most cases thus resulting in worsening health status and most cases, death. The number of radiologists in Ghana is woefully inadequate thereby hindering the achievement of equitable and quality healthcare delivery, [7]. Teleradiology would therefore avoid patient travel, significantly reduce out-of-pocket expenses and provide a timely and accurate diagnosis, [8].

The immense benefits of teleradiology were also seen especially during the COVID-19 pandemic with several countries adopting its use to address the severe impediments the pandemic created in healthcare delivery, [9]. With the diagnosis and treatment of the COVID-19 virus heavily dependent on radiological services such as diagnostic X-ray and computed tomography scans etc., teleradiology played a major role in maintaining the continuity of health services to patients by providing an effective, convenient, and safer mode of provision of radiological interpretations and eventual diagnosis, [9,10].

Studies have investigated the implementation of teleradiology, especially in specific African countries such as Ghana, South Africa, and Kenya, [9, 11, 12]. Including article reviews on the implementation of teleradiology in Africa, [8]. This paper also aims at examining teleradiology in Africa by purposively using countries from East, West, Southern, and Central Africa. The use of articles from diverse countries makes this paper unique from other publications such as that of Tahir et al, who only focused on Nigeria. Furthermore, this study fills the gap of Ewing & Holmes, who used only six articles from six African countries to make conclusions on the experiences of teleradiology in Africa, [13]. Moreover, these article reviews have not focused holistically on the various teleradiology modalities used by these countries and the successes or failures encountered during the implementation and use of teleradiology in these African countries.

Therefore, this study aims to review the extent of the implementation of teleradiology in these countries, their challenges, and the teleradiology modalities adopted. At the end of this study, we believe we will able to provide an evidence-based structure for policymakers to work with, in the quest for the widespread adoption of teleradiology in Africa.

In this systematic review, we looked closely at teleradiology implementation and adoption in five African countries namely Ghana, Kenya, South Africa, Egypt, and Morocco. These countries were purposively selected based on their geographical locations (South, West, East, and Northern Africa) which gives a fair representation of Africa in this study. In the selected countries, the literature supports the general growing discrepancies between patient needs and the number of available radiologists thus hammering on the need for teleradiology, [5,7].

## 1.2 Methodology

### 1.2.1 Research Process

First, relevant databases were picked, including one that is unique to telemedicine (i.e., Pub Med); three were evaluated for their multidisciplinary perspective (i.e., google scholar, and Web of Science and general Google search); In furtherance, online databases such as EBSCO, Scopus, and Science Direct were used to search for relevant literature. To develop a list of keywords for this paper and include relevant synonyms, the keywords identified in some published papers were relied upon, that is thesaurus terms found in various databases. In addition, keywords appearing frequently in articles dealing with telemedicine were identified. After trialing various word combinations, the following words were selected for the word “telemedicine” OR telehealth” “teleradiology” OR “mobile teleradiology”) AND “South Africa” OR “Morocco” OR “Kenya” OR “Egypt” OR “Ghana” AND “COVID-19” OR “coronavirus disease” OR “rural areas”. Again, about related terms, words like Again, about related terms, words like “artificial intelligence” OR “deep learning” OR “chest film” OR “X-rays” were used.

### 1.2.2 Research Strategy

General terms like “telemedicine*” and “telehealth*” were used as keywords to ensure a thorough search, which led to a sizable percentage of the retrieved articles not being related to teleradiology. To weed out studies that weren’t relevant, the initial round of publications was evaluated based on their titles and abstracts. Using the backward and forward snowballing strategy, 15 articles were found to help in the search [14]. To find publications that cited the retrieved papers, cross-referencing was also done for backward snowballing while Google Scholar was used for forward snowballing. The publications that had been found were evaluated using the “title and abstract review criteria.”

Through a full-text analysis of the articles, the analysis’s final sample was chosen. Nguyen and colleagues devised two sets of criteria: topic relevance and quality, which were used to evaluate each publication. According to Nguyen et al.’s quality standards, the study reviewed the articles for their relevance to the research goals and rated them on the grounds of theory, methodology, and techniques, analysis, relevance, and contribution. An article had to meet the basic level of quality standards across the board as well as at least one of the Level 3 quality parameters in order to be considered for further analysis. The search turned up 105 papers, and after 65 of them had duplicates deleted, 40 publications remained. 19 publications were chosen for this study after using the inclusion criterion of a publication date between 2015 and 2022.

### 1.2.3 Quality Validation

The quality of an SLR refers to the extent to which the study minimizes bias and maximizes internal and external validity. To ensure the reliability of inclusion and exclusion decisions, this paper applied the test-retest approach. Thus, this paper re-evaluated a random sample of selected articles after the initial analysis and validated coherence decisions based on established criteria [15]. Inclusion criteria were as follows: (a) studies related to the development and use of teleradiology in the African continent. To do these five different countries namely Ghana, Egypt, Morocco, Kenya, and South Africa were purposively selected. These countries were purposively selected because they are either in West Africa (Ghana), North Africa (Morocco, Egypt), East Africa (Kenya), and South Africa (South Africa).

Furthermore, the articles needed for this study were found in these countries. Two countries were included from North Africa to complement the maximum number of articles needed for this study. Whereas only one article was found in Ghana no other country in West Africa had articles on teleradiology based on the objectives of this study (b) articles that included the application of artificial intelligence in radiological setups to improve teleradiology or radiological consultation or interpretation among the five different countries. Studies differed to some extent in their definition or inclusion of various artificial intelligence or deep and machine learning techniques. Papers that were less than 8 years as at the time of writing this paper were excluded from the study. therefore, the papers used in this study ranged from (2015-2022). The data collected from each article included the name of the author, year of publication, the country in which the study originated, study design or method, title, objective, result and findings, and gap analysis or limitations of each study. Data from the final selected resources are charted in Table 1 below.

**Table 1:** Literature review of related articles on teleradiology in Kenya, Ghana, Egypt, Morocco, and South Africa.

| S/N | Name of Author(s) | Title | Objective(s) | Methods | Results | Gap analysis |
| --- | --- | --- | --- | --- | --- | --- |
| **SOUTH AFRICA** |  |  |  |  |  |  |
| 1. | Essop, H. (2018). [11] | Exploring the views of teleradiology end-users regarding its usage and impact on rural health services in Dr. RSM district North-West Province. | The purpose of the study was to investigate and characterize the views of teleradiology end users regarding the use of teleradiology for CT scans and the impacts it has on the delivery of healthcare services. | Qualitative research using a focus group discussion with radiology end users | 1. In the Dr. RSM district, teleradiology is used to perform CT scans.<br>2. The end users of teleradiology respect one another's professional identities.<br>3. End users of onsite teleradiology desire to upgrade their skills, and d) teleradiology infrastructure has to be strengthened. | Instead of drawing broad conclusions, this study sought to understand more about end users' perspectives. This was attributed to a number of provinces, such as Limpopo, the Northern Cape, and Mpumalanga, not making any comments. The study was further limited to CT scans and the necessity of radiologists providing direction or interpreting the CT images. |
| 2. | Ewing, B., & Holmes, D. (2022). [13] | Evaluation of Current and Former Teleradiology Systems in Africa: A Review. *Annals of Global Health*, 88(1). | To examine Teleradiology development in South Africa. | Descriptive | According to the report, tele-radiology and telemedicine advancements in Africa, particularly South Africa, were frequently sparse and modest pilot-type operations before the late 1990s. The first significant development in tele-radiology was also made in 1998 by the Department of Health (DOH) in South Africa. The DOH in | 1. The research had a relatively tiny sample size.<br>2. Tele-radiological services are used in modest and sporadic numbers. |
|  |  |  |  |  | South Africa planned to set up a national telemedicine system in an effort to enhance primary healthcare services there. |  |
|  | Essop, H., & Kekana, M. (2019). [5] | The Need for Improved Telecommunication and Collaborative Practice Among Teleradiology End Users in a Rural District of South Africa. Journal of Radiology Nursing, 38(4), 281-285. | This study shows how interactions between these medical personnel in a rural South African area affect the quality of the teleradiology service rendered. | A qualitative technique was used, and the study design was exploratory descriptive. | There are significant strains in the interprofessional relationships between all end users as a result of miscommunication. end users at the teleradiology site lack support from the remote radiology service provider, who must provide guidance and educational support. | The primary drawback of this study is that it only addresses communication issues in one teleradiology program. |
| 3 | Ntja et al (2022). [22] | Diagnostic accuracy and reliability of smartphone-captured radiologic images communicated via WhatsApp® | The study's goal was to evaluate the diagnostic precision and dependability of radiologic pictures obtained using a smartphone to those obtained using picture archiving and communication systems (PACS). | From the Pelonomi Tertiary Hospital's PACS system, radiographs from June 2018 to July 2019 were chosen for a cross-sectional study. | A sample of 135 X-rays that represented typical emergency situations was chosen. PACS accuracy was generally greater than smartphone accuracy for all registrars. All of the Kappa values suggested that there was fair to moderate agreement between smartphone and PACS diagnosis. | The screen resolution of the phone used may be a reason for the significant value obtained when the accuracy of images sent via smartphone and PACS were compared. Another reason may be the size of the screen. Bigger screen sizes yield better results since u can view from different angles. |
| 4. | Mbunge et al (2022). [16] | Virtual healthcare services and digital health technologies deployed during the coronavirus disease 2019 | To list the digital health technologies and virtual healthcare | Observational | This study revealed South Africa adopted various virtual healthcare services such as Telemedicine during COVID- | Lack of image interpretation training for physicians or other health professionals. |
|  |  | (COVID-19) pandemic in South Africa | services used in South Africa during the coronavirus disease 2019 (COVID-19) and the difficulties using them. |  | 19. In the implementation of Teleradiology, it was noted that disparities in radiology research and infrastructure have been reported in South Africa. | Limited funding and insufficient healthcare workers in the rural areas. Lack of maintenance culture to ensure the effectiveness of teleradiological systems such as PACS. |
| **EGYPT** |  |  |  |  |  |  |
| 5. | Abodahab et al. (2021). [19] | Implementations of PACS and Teleradiology Systems at Sohag University Hospital | To assess and improve the current usage of PACS and Teleradiology systems in Sohag University Hospital generally, and the Radiology Department particularly, as well as acquired experiences with them. | An experimental study to evaluate the use of Millensys PACS "workspace version 5.0.0.1496" from 15 November 2015 to 1 June 2017 inside 18 months | 57453 instances in total were supplied via PACS over the course of the study. 7.50% of them were CT cases and 3.03% were MRI cases, while 89.47% of them were from the CR. There has been a decline in the number of cases forwarded to PACS in several months. Only 8.7 percent of sent CT cases and 0 percent of sent MRI cases were verified over the course of the study. Only 4.9% of sent CT cases and 17% of sent CT cases were reported on our PACS. representing 0% of MRI cases and 0.01% of CT cases. The overall PACS cost savings for Sohag University Hospital throughout the course of the project came to 1550000 EGP. | This study although very revealing, had a much better outcome or result was hindered by insufficient computers connected to PACS and low maintenance of fixed IP for internet access. |
| 6. | Alfarghaly et al. (2021). [23] | Automated radiology report generation using conditioned transformers. | This study suggests a deep learning model to produce radiologist reports automatically given a chest X-ray picture from the publicly available IU-Xray dataset. | (1) Develop a pre-trained Chexnet to better anticipate certain tags from the picture. (2) Using the pre-trained embeddings of the expected tag, determine the -weighted semantic features. (3) To produce the complete medical reports, pre-train a GPT2 model on the visual and semantic characteristics. | The outcomes demonstrated that the CDGPT2 model outperformed other non-hierarchical models in terms of average Bleu score for word-overlap measures. The reason for the decline in Bleu-3 and Bleu-4 using this approach is that RNCM had a greater bleu average due to their high Bleu-1 score, but they only forecasted tags to provide a restricted controlled report. To condition a pre-trained DistilGPT2 on visual and semantic aspects and produce full-text reports for chest X-Ray pictures in the nation, a new conditioning approach was introduced. | The dataset size used was small thereby limiting the model's generalization and implementation. There was less funding to expand the study. |
| 7. | Mohammed et al. (2020) [24] | The battle against Covid-19: the experience of an Egyptian radiology department in a university setting. | This paper intends to detail the different procedures carried out by a radiology department in an | Observational | Before the COVID-19 pandemic, the Egyptian radiology department successfully implemented home access to picture archiving and communication systems (PACS) (Fuji synapse) | The hospital occasionally relies heavily on the actual presence of patients for one-on-one contacts between radiologists and residents for |
|  |  |  | educational institution in a resource-limited nation during the COVID-19 crisis, offering insights into the employed tactics in other institutions in developed nations. |  | to safeguard patients, protect radiology faculty staff, and conduct remote radiological consultation and examination. | instructional reasons, between the referring doctors and technicians, hence there was very little usage of the PACS. Additionally, it was difficult for radiologists to ensure a dependable and stable internet connection, and it was difficult to purchase workstations for home PACS due to a lack of resources. These factors all reduced the effectiveness of the PACS implementation in the radiology department of the hospital setting. |
| 8 | Ajlan et al (2021). [25] | Detectability of cerebral chronic white matter microangiopathy on CT compared to MRI: A teleradiology study. | WM microangiopathy's capacity to be detected on CT as opposed to MRI using a subjective visual method. | In a private Jeddah hospital in 2020–2021, a retrospective chart review was conducted. | The juxtacortical area is the primary location of smaller lesions that were more frequently overlooked on CT scans.<br>20% of all WM locations showed CT WM alterations, whereas 8.6% of each of the deep and juxtacortical WM regions did as well. | Untreated elderly lacunar infarcts are regarded as a component of chronic white matter small vessel disease. Acute ischemic or hemorrhagic infarctions and the relationship between those acute observations and chronic WM microangiopathy are another significant imaging result that was not assessed in this investigation. |
| **MOROCCO** |  |  |  |  |  |  |
| 9. | Benbrahim et al (2020). [26] | Deep Transfer Learning with Apache Spark to Detect COVID-19 in Chest X-ray Images | Convolutional Neural Networks (CNN) based on the pre-trained models InceptionV3 and ResNet50 will be used to create a system based on Deep Transfer Learning (DTL) within the Apache Spark big data framework. | Experimental | For the purpose of identifying COVID-19 in chest X-ray pictures, a strategy built on deep transfer learning and utilizing CNN based on Inception V3 and ResNet 50 with Apache Spark has been presented. For Inception V3 and ResNet 50 models, the developed model attained high accuracy of 99.01% and 98.03%, respectively. This model complied with the logistic regression algorithm and provided significant results for accurately detecting COVID-19 in chest X-ray (CXR) images, which can be used by health professionals in both rural and urban areas. | For the identification of COVID-19 in chest X-ray pictures, a deep transfer learning technique based on CNN built on Inception V3 and ResNet 50 with Apache Spark has been presented. For Inception V3 and ResNet 50 models, the developed model attained high accuracy of 99.01% and 98.03%, respectively. This model complied with the logistic regression algorithm and produced significant findings for accurately detecting COVID-19 in chest X-ray (CXR) images, which can be used by health professionals in both rural and urban areas. |
| 10. | Marti-Bonmati et al (2022). [27] | Considerations for artificial intelligence clinical impact in oncologic imaging: an AI4HI position paper. | The primary general goal of AI-based studies involving cancer imaging data is to provide decision-support tools from clinical-molecular data and standard-of-care images by giving doctors estimates or predictions of disease aggressiveness, expected treatment response, and final clinical outcome. | Descriptive | The inter-disciplinary African community of active contributors to AI for radiology and imaging applications was formed, maintained, and expanded through the AFRICA initiative. It was the creation of dependable, adaptable AI solutions to real-world healthcare issues unique to Africa. | Most African nations, including Morocco in this study, have extremely limited access to resources for basic medical imaging and image analysis. |
| 11. | Lafraxo, S., & El Ansari, M. (2021). [28] | CoviNet: automated COVID-19 detection from X-rays using deep learning techniques | The primary objective of this study is to provide a practical foundation for the COVID-19 automated diagnostic. | Experimental | With an accuracy of 98.6390 and 95.7790 for binary classification and 95.7790 for multiclass classification, the coriNet system was created to be utilized for both categorization and differentiation. Health professionals in Morocco can employ CXR pictures into COVID-19, normal or effusion, in both urban and rural locations. | Less clinical datasets were used which could not better evaluate the effectiveness of the model thus hindering its use in early detection of COVID-19 in health centers as in various teleradiological sites. Lack of funding to expand the study. |
| 12. | Marwan, M., Kartit, A., & Ouahmane, H. (2018). [29] | A cloud-based framework to secure medical image processing. | The fundamental purpose of this research is to present an innovative way for safe cloud-based medical image processing. | Experimental | In addition to proposing a solution based on a generic algorithm and segmentation, a generic framework was created to safely process patient data. A new trusted entity, or CloudSec module, was added to an existing traditional cloud architecture. The simulation results demonstrated that the suggestion was a sufficient method for enhancing image analysis utilizing public cloud computing and that it considerably enhanced both data security and performance. | It takes a lot of time, which has a negative impact on the radiological system's quality of Service (QoS). Image data is not sufficiently safeguarded by the service-oriented architecture (SOA) against unreliable cloud providers. Lack of funding to expand the study thereby hindering the effectiveness of tele-radiological implementation and cloud computing. |
| 13. | Boufarasse et al (2020). [10] | Teleradiology and AI as Solution to Overcome the COVID-19 Pandemic Impact during the Lockdowns in Africa. | The study's objective is to show how teleradiology and artificial intelligence can be used in a pandemic. | Exploratory study | To protect professional and patient safety and increase production during the COVID-19 crisis, teleradiology, artificial intelligence, and teleworking are urgently suggested in Africa. | It did not provide guidance on how to keep cybersecurity at its best. |
| **KENYA** |  |  |  |  |  |  |
| 14 | Kavoji, S. H. (2021). [12] | User Acceptance of Teleradiology in Kenya a Case Study of Two Teleradiology Centers in Nairobi County | A Case Study of Two Centers in the County of Nairobi, Kenya, Regarding User | Cross-sectional survey at two teleradiology | The study's data findings revealed that performance expectations had a regression coefficient of $=0.017$ and a significance value of $p=0.011$, | Insufficient data was chosen for the study since just two teleradiology facilities in Nairobi have used |
|  |  |  | Acceptance of Teleradiology. | centers in Nairobi. | indicating that it substantially and positively influenced teleradiology acceptance. The regression coefficient and significance values for the effort expectation, technological experience, enabling circumstances, and training adequacy all indicated that these factors substantially and favorably affected teleradiology adoption. | the technique. Instead of 60 participants, only about 45 were included in the study's sample size. |
| 15. | Mwogi et al (2018). [17] | A scalable low-cost multi-hospital teleradiology architecture in Kenya | Give an account of the creation in Kenya of a multi-hospital, affordable, scalable teleradiology architecture. | A January–December 2017 experimental research conducted among four private clinics in western Kenya. | Launched in January 2017, the teleradiology system. There were 1,335 CT scans, 150 MRIs, and 80 X-rays (CR studies) among the investigations that were received and analyzed from January to December 2017. Within three ($3 \pm 1.4$) minutes of the transfer's start, all of the studies had been completely transferred to the main DICOM server and were ready for radiologists to report on right away. The lossless compression method was applied to all investigations. | In this work, multimodal security strategies to safeguard patient-level data in teleradiology systems were not investigated. |
| 16. | Larrison, M. (2016). [30] | Teleradiology Service for Mission Hospitals: Initial Experiences. | To examine Experiences with teleradiology in Soddo Christian | A cross-sectional study involving | The fact that these radiologists take part in teleradiology throughout the year has also allowed them to keep in touch | This research compared the teleradiology systems at Tenwek Hospital and Soddo |
|  |  |  | Hospital and Tenwek Hospital | Soddo Christian Hospital and Tenwek Hospital | with Tenwek on a regular basis. | Christian Hospital, focusing on access as the point of comparison. Other hospitals should be considered in future studies. |
|  | Essop, H., & Kekana, M. (2020). [5] | The experiences of teleradiology end users regarding role extension in a rural district of the North West province: A qualitative analysis. | To investigate how the experiences of the end users within this setting affect the service. | This was a qualitative, exploratory, descriptive study. in North West Province, South Africa | Radiographers and referring doctors are filling out additional duties at the teleradiology site that are not covered by the teleradiology service-level agreement (SLA) and for which they feel ill-prepared. In order to comprehend the difficulties faced by health workers in these remote locations, they also believed that private radiologists required training in interprofessional teamwork. | The results of this study cannot be applied to other contexts since they are exclusive to this primary healthcare environment. To discover particular difficulties, we advise doing more investigations in teleradiology situations. |
| 17. | Vinayak, S. (2017). [31] | Training midwives to perform basic obstetric pocus in rural areas using a tablet platform and mobile phone transmission technology | This investigation sought to assess a number of procedures, including the precision of the images and reports produced by midwives, the functionality of a tablet-sized ultrasound | Cross-sectional involving midwives in rural locations where POCUS was available for the first time | From the conclusion of the scan to the scan confirmation of the report, the total turnaround time was around 35 minutes. The mobile/cell phone model, CCC system (Philips Medical Solutions), or other difficulties were not experienced. There was no difference in transmission speed, and no image quality reduction was discovered. The | Only three midwives were evaluated making the sample size of the study very small. There was a risk of selection bias in the study and an experienced sonographer was not used in assessing the training aspect of the study. |
|  |  |  | scanner, training of midwives to perform ultrasounds, teleradiology solution image transmissions via the internet, review of images by a radiologist, communication between the midwife and radiologist, use of this technique to identify high-risk patients, and improvement of education and teleradiology model. |  | radiologist had quick internet access to the shop images. |  |
| 18. | Vinayak et al (2017). [18] | Training Midwives to Perform Basic Obstetric Point-of-Care Ultrasound in Rural Areas Using a Tablet Platform and Mobile Phone Transmission Technology-A WFUMB COE Project | To assess the precision of pictures and reports provided by skilled midwives performing basic obstetric ultrasonography exams at our satellite sites. | Prospective cross-sectional study | The midwives did 271 ultrasound scans in all. Two highly competent radiologists from AKUHN examined all of the data, including the pictures and related measures, and found that everything complied with the methodology's accepted norms and criteria. The midwives' scans' measurements and photos had a 99.63% accuracy rate. | While highly successful, this study is on a small scale. It will take careful planning and resource acquisition to test scalability, which has not yet been done. |
|  | Kamau et al (2018). [32] | Towards A More Secure Web Based Tele Radiology System: A Steganographic Approach. | The major goal of this study was to look into the specifications for a safe LSB-based steganographic technique that increases the classic LSB technique's levels of imperceptibility while being flexible enough to meet the demands of a web-based teleradiology infrastructure. | Utilizing the "learning through building artifact construction" research methodology, Design Science Research (DSR). | The outcomes showed that the suggested strategy may reduce MSE ratios by up to 0.4 and enhance Peak Signal Noise Ratio (PSNR) by 1 to 2.6 decibels (dB). | The study is prone to the limitations of The ELSB method |
|  | Larrison et al (2016). [33] | Teleradiology service for mission hospitals: initial experiences in Ethiopia and Kenya. | To examine teleradiology practices in mission hospitals | Observational study of mission hospitals in Kenya and Ethiopia | These radiologists' year-round participation in teleradiology has also made it possible for them to maintain regular contact with Tenwek. | The teleradiology systems of Soddo Christian Hospital and Tenwek Hospital share this access. |
| **GHANA** |  |  |  |  |  |  |
| **19** | Edzie et al (2020). [7] | Application of information and communication technology in radiological practices: a cross-sectional study among radiologists in Ghana. | 1. To determine how ICT use affects radiologists' ability to do work.<br>2. To ascertain the imaging | During the Ghana Association of Radiologists' annual general meeting, a cross-sectional | All of the radiologist present were given a total of 46 questionnaires, and all (100%) of them were returned and documented. Radiologists had access to PACs, EPR, RIS, and tele-radiology, respectively, for | There is a limited sample size. Unreliable internet, poor network accessibility, frequent power outages, computer system failures, and a lack of |
|  |  |  | techniques offered by Ghanaian radiological practitioners.<br>3. To find out if ICT had an impact on how long radiological reports took.<br>4. To discover the extent of social media use in radiology offices. | study involving 46 willing radiologists was carried out from May 16 to May 18 of this year. | 63.0%, 39.1%, 39.1%, and 13.0% of the radiologists. The respondents were conversant with PACs, EPR, and RIS to varying degrees (78.3%, 54.3%, and 52.2%, respectively). Additionally, only 32.6% of respondents in teleradiology reported having experience with software. At a 95% confidence level, an association study to investigate the connection between years of experience and availability of ICT tools found no correlation. 41.3% of the radiologists did not utilize DICOM in their practices, compared to 59.7% of them. The majority of responders (91.3% and 73.9%, respectively) did not utilize the MicroDicom and AMIDE tools. Of the 46 participants (radiologists), 69.6% strongly agreed and 30.45% agreed that conducting online research aided in the gathering of data for report writing. | technical know-how are a few of the difficulties encountered, which contribute to the limited availability and low familiarity with the use of tele-radiology in the majority of the country. |

## 1.3 Descriptive Analysis of the Literature

The main methodologies were identified in the literature on the teleradiology subject, qualitative, quantitative, and experimental research approaches. In the overall sample, methodologies range from about 50% purely quantitative to 15% purely qualitative and 35% experimental. Additionally, in terms of data collection instruments, surveys make up the largest number of empirical studies, followed by experimentations, and interviews. The Theory identified was the technology acceptance theory. This idea, which holds that two elements affect whether a computer system will be accepted by its potential users, was employed in the study of, [7]. These are perceived applicability and perceived usability, [7]. This theory is useful to this type of study because teleradiology implementation involves the use of technology. According to theory people, both patients and doctors will adopt teleradiology practices if perceive it to be useful and it is easy to use. The highest number of articles were obtained in the order Kenya, Morocco, South Africa, and Ghana.

## 1.4 Discussion of Objectives

### 1.4.1 Implementation of Teleradiology and modalities adopted

It was found that despite the low adoption of teleradiology before the year 2019, there was a spike in the applications of teleradiology as it was among the digital healthcare services adopted during the COVID-19 pandemic which was during and after the year 2019 in so many countries. This was because teleradiology was among the digital healthcare services adopted during the pandemic. According to [16] among the other modalities of telemedicine actively used in South Africa, teleradiology recorded significant healthcare benefits in the prompt diagnosis of disorders, particularly concerning lung health, [16].

Kenya has seen an appreciable number of adoptions and works on teleradiology over the years. For instance, in 2018, [17] explored a study on a low-cost Teleradiology architecture where the system explored full-scale modality applications of digital X-ray, MRI, and CT scan, [17]. This study saw tremendous success with image transmission and reporting timelines across all three modalities. Also, Vinayak et al, have reported the successful deployment of teleradiology with the utilization of Point-of-Care Ultrasound (POCUS) by midwives. It was estimated that the total flow turnaround time was 25 minutes (from patient presentation to validated report), which was a very significant amount of time, particularly when caring for high-risk patients in distant healthcare settings, [18]. For a successful teleradiological implementation, human acceptance plays a vital role so, [12] in 2021, thoroughly reported on user resistance as part of measures in teleradiology adoption, [12]. The study carried out in two teleradiology centers in Nairobi concluded that there was an appreciable acceptance rate by the users. The overall user approval of teleradiology systems, however, was shown to be significantly influenced by training adequacy as it was discovered to be a recurrent answer from respondents.

Studies in Egypt also saw the implementation of teleradiology systems employing CT scans and MRI modalities in their application. A large number of CT and MRI images were sent through Picture Archiving and Communication Systems (PACS) for reporting and interpretation, and the adoption was largely effective, according to, [19]. There were no issues noted with transmission through the PACS system,[19].

Nearly all of the published articles and studies noted teleradiology’s significant contribution in some way. Almost all studies or articles that looked at satisfaction across the five nations (Ghana, Egypt, Morocco, Kenya, and South Africa) revealed that doctors and patients had generally positive opinions of the teleradiology programs that had been put in place there.

### 1.4.2 Challenges in the Implementation and Use of Teleradiology

The publications and research under examination make it clear that, while having significant potential, the teleradiology system is not being used to its fullest extent. Very little work on teleradiology is reported across Africa specifically in the five countries of focus, South Africa, Egypt, Kenya, Morocco, and Ghana from the year (2015-2022). The few articles identified have shown some challenges of diverse formats regarding the implementation and use of teleradiology. These challenges have been classified as training challenges, Human resource challenges, technological barriers, and policy issues.

#### 1.4.2.1 Training

There is a paucity of training on the application and benefits of teleradiology for the parties engaged in Kenya, Ghana, and Egypt, [13], [17], [20]. In addition, it was found that none of the countries that had introduced teleradiology had enough in-service training for its medical professionals. Therefore, radiologists and other stakeholders need to have the appropriate training to assist in mitigating some of the issues that are linked to the use, [5] stated that a lack of training and change management was another factor that contributed to the low utilization of teleradiology in Africa, [5]. It is essential to receive training on how to make use of these teleradiology devices, particularly for radiologists. Despite this, it is extremely vital to provide patients with at least some level of education regarding teleradiology. This is because even if some medical professionals could be resistant to incorporating new technologies into their practices, ongoing education will assist mold their abilities to do so. Education of patients on the significance of and benefits associated with the utilization of teleradiology may assist improve their preference for teleradiology practices, which, in the long run, may persuade hesitant medical professionals to begin utilizing teleradiology. Therefore, once a nation has decided to implement teleradiology, systems need to be put in place to ensure that radiologists and other stakeholders participating in teleradiological practices receive regular training. Teleradiological practices

#### 1.4.2.2 Human Resources

In addition, the results of our investigations suggest that the majority of the medical facilities make use of teleradiology, which falls under the category of extramural teleradiology and involves radiologists who are not permanent employees of the medical facilities in question. This has resulted in a breakdown in communication between the referring physicians and the on-site radiologists, which has been compounded by the lack of a Service Level Agreement (SLA) or the ineffective implementation of the policy. According to Britton et al, some of the drawbacks of extramural teleradiology include the following: a disruption in the interaction between referring physicians and radiologists; a reduction in the leadership of local radiologists, [20]. These restrictions are solvable problems that can be remedied by evaluating the SLA and enforcing it more strictly. This alludes to the fact that even though teleradiology is an efficient technology that will benefit African countries, the challenge of the shortage of radiologists must be mitigated, [19]. Due to this, these countries have still not fully adopted the use of teleradiology within the context of their healthcare delivery. As evident in the use of teleradiology in only a few parts of the country or only in major hospitals in urban settings and many hospitals in rural settings.

#### 1.4.2.3 Technological Barriers

Despite the overwhelming benefits, the full implementation of teleradiology was hampered by barriers caused by unavailable technology. End-user experiences have shown that there are hinges in the teleradiology architecture that has been used in the few nations that have a written paper on the architecture of the teleradiology system that has been implemented. Radiographers and referring doctors were undertaking expanded activities beyond of their authorized scope, and they also felt ill-equipped to carry out their duties effectively, according to the experiences of teleradiology end users, [5]. This was a problem for the end users because they were exceeding the allowable scope of their responsibilities. The results of this study are consistent with those of Ali et al, who proposed that, in order for teleradiology to be more successful given the enormous volumes of data produced, improving the storage capabilities of the servers used for teleradiology data must be a top priority, [21]. These technological hurdles provide a significant risk to the prospects of any nation’s teleradiology architecture being successful when it is put into place. This is due to the fact that teleradiology is driven by technology. Therefore, if there are challenges to the same technology that was built, it might potentially have a catastrophic effect. The lack of financial means to purchase the necessary technology for teleradiological practices is the primary factor contributing to the existence of these technological hurdles. Particularly in Africa, where the full potential of technological advancement has not yet been realized. Africa is completely reliant on the technologies that the Western world pioneered. Hence without the initiatives of Western donors to implement teleradiology in certain parts of Africa, it is most likely that there will be no government initiative to do so. As a result, the cost of such technology rises because of the fluctuating exchange rates required to acquire various pieces of technological equipment denominated in a foreign currency.

#### 1.4.2.4 Policy

It is impossible to assess the effectiveness of a policy or program guiding teleradiology in Ghana since there is no strategic retention policy that is suited to rural radiography practice, [7]. The findings from Ghana can, to a significant degree, be generalized to apply to other countries in Africa, even though other research did not demonstrate a distinct absence of policy on the usage of teleradiology. This is due to the fact that technology does not significantly affect how health care services are delivered in Africa; hence, there are no rules that set limits on how much technology should be used by various health institutions in Africa. The manner in which those technologies will be utilized to guarantee equal delivery across the country over time, in addition to any other regulations that are particularly geared toward the utilization of teleradiology. Research that strives to understand the attitudes and experiences of radiographers who operate in rural regions of sub-Saharan African countries is thus required. The results of this study might then be applied to the creation and implementation of policies.

## 1.5 Conclusion

Teleradiology is the process of transmitting radiological patient pictures (including X-rays, CT scans, and MRIs) over long distances in order to share these studies with other doctors for consultation or interpretation. This is one of the most well-established, prosperous, and popular subspecialties in clinical telemedicine. This study provides evidence that advances in hardware and information technology have made it possible to push the field of teleradiology even farther and to expand the market for mobile teleradiology. picture quality, transmission speed, and picture compression were once considered to be technological barriers that modern smartphones and mobile networks have surmounted. However, there are still problems like a lack of sufficient and dependable local connectivity and a lack of electricity.

It additionally illustrates that teleradiology, including mobile teleradiology, is not only simple, useful, and appropriate for assisting patients and healthcare providers with routine X-rays in rural and remote areas, but also that it is dreadfully underutilized in Ghana and Kenya, despite the fact that there is not enough regional evidence to adequately support this. Despite this, research has shown that teleradiology may improve patient care and healthcare delivery (for example, by increasing the referring doctor’s diagnostic accuracy) and is a practical alternative in places where there are few or no radiologists. The nearly ubiquitous usage of mobile teleradiology and teleradiology for imaging procedures that are considered routine elsewhere is a reflection of the quickly expanding domains of clinical practice, service delivery, and technology.

According to the studies that have been done, the lack of advanced teleradiology in the African region restricts patients’ access to the most fundamental of radiological services. The introduction of teleradiology programs in the nations indicated above has an immediate requirement for extensive published research that is designed to create high-quality and evidence-based recommendations for best practices. This would be in line with the assistance the World Health Organization (WHO) has given to mobile health, the achievement of universal health coverage (UHC), and the Sustainable Development Goals (SDGs) set forth by the UN, notably objective 3: Good Health and Well-Being.

## 1.6 Limitations of this systematic review and Direction for further research

The study has several constraints. Only two citation databases, PubMed and publish or perish, as well as Google Scholar, were searched. Even though they were discovered, not all of the other mobile-teleradiology-related problems were included. The level and quantity of information supplied during consultations, data security during transmission, data storage and record keeping, and patient identity, for example, were not recorded. These elements must be taken into account while developing any treatment program. Additionally, there was a lack of a thorough charting of teleradiology-specific issues like camera and screen resolution, file format, and the use of RIS and PACS. Therefore, further studies should comb other databases and also focus on the diverse issues that are present in articles but not discussed in this study. Further studies should comb the literature to identify whether the gaps identified by individual studies were addressed by other empirical studies in these countries or other countries that were not used in this study. Most of the articles were obtained from an East African country (Kenya) whereas the least number of articles were from a West African country (Ghana). In West Africa, there was only one article found on teleradiology. Therefore, scholars can focus on examining various aspects of the implementation of teleradiology in West Africa.

## 1.7 Recommendation for practice

Based on the fact that decision-makers and policymakers require local evidence to make choices, and based on the findings of this analysis, it is suggested that:

- A select number of well-built and detailed studies should be conducted to demonstrate the usefulness and cost-effectiveness of mobile teleradiology against traditional radiology, as well as to give clear evidence-based advice for deploying mobile teleradiology programs.
- Every solution has an opportunity cost and must meet a real and urgent need, thus any rural or distant healthcare institution contemplating mobile teleradiology must do a local needs assessment to verify predicted feasibility.
- Once sustainability has been established, mobile teleradiology should gradually be implemented with the necessary guidance and assistance.
- In-service training must be continuously done to update staff on the current mode of operations of teleradiology.
- Servers used must have huge capacities to accommodate the data generated.
- There should be a stricter enforcement and clearer definitions of service level agreements to ensure effective communications between physicians and extramural radiologists.

## Data Availability

All data produced in the present study are available upon reasonable request to the authors

## Acknowledgement

We would like to express our utmost appreciation to our supervisor, Dr. E.O Osei for his significant guidance and support during the research process. His insights were valuable in determining the scope and direction of our work.

We would also want to thank our colleagues from Kwame Nkrumah University of Science and Technology for their recommendations and support during this research. These contributions were pertinent to the research topic and drew pertinent findings.

Finally, we would like to thank the reviewers for their thorough analysis of this manuscript.

## Funding Statement

For this investigation, the authors got no financial support.

## Compliance

In the course of this systematic review, no animal or human data were used.

## Conflicts of Interest

The study’s authors all certify that they have no competing interests to disclose.

## Authors Contribution

**Conceptualization:** All authors contributed to developing the study concept.

**Methodology:** All authors contributed to developing the study methodology.

**Resources:** All authors participated in providing resources in terms of searching for articles fitting the inclusion criteria of our study.

**Data Curation:** All authors were equally involved in data or resource curation. However, MG performed the final confirmatory data curation.

**Visualization:** MG performed all data visualization.

**Supervision:** LAA performed supervision during the research process.

**Project Management:** LAA performed the administration and management of this study project.

## Notes

### Competing Interest Statement

The authors have declared no competing interest.

### Funding Statement

This study did not receive any funding

